# Positive Running: a gender-transformative physical activity intervention to overcome intersectional barriers among adolescents with perinatally acquired HIV in India

**DOI:** 10.64898/2026.02.17.26346488

**Authors:** Siddha Sannigrahi, Kacie Filian, Babu Seenappa, Hrishikesh Sathyamoorthy, Suhas Reddy, Meghana Gowda, Jeevitha, Ramu Sanju, Sahana Papanna, SK Satish Kumar, Michael Babu Raj, Lakshmi Ganapathi, Anita Shet

**Affiliations:** Johns Hopkins Bloomberg School of Public Health; RISHI Foundation; YRG CARE: YR Gaitonde Centre for AIDS Research and Education; Boston Children’s Hospital

## Abstract

**Background:** Adolescents with perinatally acquired HIV in India experience a high burden of stigma and mental health distress alongside gendered social constraints that limit participation in supportive programs. While physical activity-based psychosocial interventions show promise for improving adolescent mental health, little is known about how gender norms and intersecting vulnerabilities shape engagement and outcomes among this population. This study examined gender-specific patterns of participation and associations with mental health in a peer-led running intervention in southern India using intersectionality and self-determination theory.

**Methods:** We conducted a convergent parallel mixed-methods evaluation between March-April 2024 among 150 adolescents and young adults with perinatally acquired HIV enrolled in a physical activity intervention (*Positive Running*) in Karnataka and Tamil Nadu. Surveys assessed sociodemographic characteristics, viral suppression, intervention adherence, and common mental health disorders using validated screening tools for depression (PHQ-9) and anxiety (GAD-7). Gender-disaggregated comparisons used Fisher’s exact tests, and logistic regression estimated prevalence odds ratios for common mental health disorders by intervention adherence. Qualitative data included four age- and gender-stratified focus group discussions (n=28) with participants, and four in-depth interviews with peer implementers. Transcripts were thematically analyzed using grounded theory and Braun & Clarke’s framework.

**Results:** Among 150 participants (100 males, 50 females; median age 17 years [IQR 15-19]), 91% were virally suppressed. Mean adherence to the intervention was 64%, with high attendance (≥65%) significantly lower among females than males (20% vs 57%, p<0.001). Overall, 59% screened positive for at least one common mental health disorder; with higher prevalence among females than males for depression (66% vs 43%, p=0.009), and for any mental health condition (72% vs 52%, p=0.022). Higher intervention adherence was associated with lower odds of common mental disorder overall (OR 0.44, 95% CI 0.23-0.85). In age-adjusted, gender-stratified analyses, this association was significant among males (aOR 0.33, 95% CI 0.14-0.75) but not among females. Qualitative findings identified gendered barriers to participation, including restrictive norms, modesty expectations, stigma toward women in sport, and limited decision-making autonomy. Self-determination theory-informed analyses highlighted how structured training, peer mentorship, and visible female role models supported autonomy, competence, and relatedness, while also revealing constraints that attenuated mental health gains for girls.

**Conclusions:** Peer-led, community-embedded physical activity interventions are feasible among adolescents and young adults with perinatally acquired HIV and may confer mental health benefits, though participation effects are gender-differentiated. Findings underscore the need for gender-responsive, autonomy-supportive program designs that address intersectional vulnerabilities to ensure equitable mental health impact, particularly for adolescent girls and young women.

## INTRODUCTION

India, which ranks third in HIV disease burden globally with an estimated 2.3 million people living with HIV, has achieved remarkable progress through comprehensive national ART initiatives, resulting in an estimated 80% decline in HIV-related mortality since 1992.^1^ India’s youth population is the largest globally; however, despite advancements in HIV care and support, adolescents account for 35% of new HIV infections annually. ^2,3^ Among groups that remain highly vulnerable are adolescents and young adults (<25 years) with perinatally acquired HIV (APHIV) who navigate a period of rapid physical, emotional, and social change, while managing a chronic, stigmatized illness.^4^ Many reach sexual maturity without comprehensive HIV knowledge, increasing risks of onward transmission if they are not virologically suppressed.^5^ The mental health burden among APHIV is substantial, with studies reporting depression rates up to 26% and generalized anxiety disorder in as many as 46%.^6–9^ These common mental disorders undermine adherence to antiretroviral therapy (ART), reduce viral suppression rates, and are linked to poorer health and social outcomes.^10–13^ Gender inequities further exacerbate these challenges. Young women living with HIV face greater stigma, less access to healthcare, and higher levels of anxiety and depression than their male peers.^14–17^ Harmful social norms often result in early school dropout, early marriage, and reduced opportunities for participation in supportive programs, thus compounding the disadvantages created by the intersection of age, gender, HIV status, and poverty.^18^

Physical activity offers a promising, underutilized pathway to improve both physical and psychological well-being in APHIV. Evidence from sub-Saharan Africa shows that sport-based interventions in youth can address mental health and social cohesion outcomes broadly, and that physical activity improves psychological status among adults living with HIV.^19,20^ However, peer-led physical activity interventions are rarely designed with a gender-specific lens, leaving adolescent girls to often face unique sociocultural constraints, including restrictive gender norms, safety concerns, and lower encouragement for sports participation compared to boys.^21^ In India’s context, culturally adapted gender-responsive physical activity interventions, particularly for APHIV remain scarce.

To address health inequities among APHIV who already experience layered stigma and vulnerability, a physical activity intervention called the *Positive Running* program was initiated in 2022, reaching over 200 young APHIV and those affected by HIV aged in the southern Indian states of Karnataka and Tamil Nadu. This study aims to investigate the gendered dynamics of motivation and participation in the *Positive Running* program among APHIV. Using Intersectionality Theory and Self-Determination Theory (SDT) as conceptual frameworks, we examined how autonomy, competence, and relatedness influence gender-specific engagement patterns and psychosocial outcomes in a population with layered vulnerabilities.

## METHODS

### The intervention: Positive Running

The *Positive Running* program is a peer-led physical and psychosocial intervention designed for adolescents and young people living with or affected by HIV. The program integrates structured physical activity with resilience-building and leadership development to promote overall health and well-being. The intervention has a weekly schedule consisting of three strength and conditioning sessions, three running sessions, and one rest day. Strength and conditioning (Monday, Wednesday, and Friday) included resistance, cardiovascular, and flexibility exercises to improve muscular strength, endurance, and mobility. Each running session targets specific performance goals: threshold runs (Tuesday) emphasize sustained moderate effort to build endurance; speed runs (Thursday) focus on short, high-intensity intervals to enhance power; and distance runs (Saturday) aim to increase aerobic capacity and mental resilience. In addition to physical training, participants attend quarterly educational camps covering topics such as nutrition, exercise physiology, hygiene, and injury prevention. These camps also emphasize resilience-building, teamwork, and goal setting to promote holistic development and psychosocial well-being. Participants also take part in 1-2 organized running events across India, providing opportunities to apply their training, build confidence, and foster a sense of community among youth living with or affected by HIV. These activities are led by youth peer implementers called Captains, who undergo training to serve as intervention implementers. They lead exercise sessions, guide participants through the program, and foster a supportive, motivating environment. Working closely with senior program leadership, Captains help maintain program consistency, encourage participation, and ensure the intervention’s overall success and sustainability.

### Community-based participatory research

Impact evaluation of the intervention was conducted through a community-based participatory research (CBPR) approach, with youth living with HIV actively engaged throughout the research process (youth investigators).^22^ Five trained youth investigators were part of the research team. Their contributions were essential in refining surveys and interview guides to improve comprehension, strengthening the study’s cultural and contextual validity, and ensuring that research activities reflected the lived realities of youth with HIV in Karnataka. All youth investigators completed Human Subjects Research certification through Johns Hopkins University and underwent a 2-day training in sensitive and ethical survey administration and qualitative research techniques.

### Study setting and participants

Between March and April 2024, we enrolled participants from childcare institutions located in Bijapur, Dakshina Kannada, and Kolar districts in Karnataka, and in Krishnagiri district in Tamil Nadu. These institutions provide residential care, psychosocial support, and educational or vocational training for children and adolescents <18 years of age living with or affected by HIV. Children in non-institutional settings and adult participants living with their respective families were enrolled from Bangalore and Bijapur. Study eligibility criteria included being (i) between 7 to <25 years of age; (ii) currently enrolled in the *Positive Running* program for at least 12 months; (iii) having documented HIV status prior to 10 years of age, or HIV-affected status (defined as having one or both parents with HIV-confirmed status).

### Study design and indicators

We employed a convergent parallel mixed-methods design incorporating quantitative and qualitative research. Following informed consent, trained youth investigators administered surveys in person, with measures in place to ensure privacy and confidentiality during data collection. These measures included conducting surveys in a private setting away from peers and staff, ensuring that responses could not be seen by others, and securely storing completed forms in locked folders accessible only to authorized research staff.

i. Survey: Survey components included sociodemographic details such as age, gender, education, and residential history. Clinical information such as key HIV-related indicators from the past 12 months, including hemoglobin, CD4 cell count, viral load, and current antiretroviral therapy (ART) regimen was obtained through participant self-report and medical records. Psychosocial health status was assessed using the 9-item, Patient Health Questionnaire (PHQ-9) and 7-item, Generalized Anxiety Disorder (GAD-7)^23,24^ to screen for depressive and anxiety symptoms. Each tool uses a 0-3 symptom-frequency scale, with PHQ-9 scores ranging from 0-27 and GAD-7 scores from 0-21. A positive screen for any depression was defined as PHQ-9 scores ≥5 (mild, moderate, or severe), while scores 0-4 was categorized as no depression. A positive screen for any anxiety was defined as GAD-7 scores ≥5 (mild, moderate, or severe), with scores 0-4 categorized as no anxiety. A counsellor was available onsite and referral systems were in place if the need arose for specialized care following administration of the mental health screening tools.
ii. Intervention adherence: The *Positive Running* intervention adherence was monitored in real-time by Captains. Daily attendance was documented on site, and was quantified as a percentage of days attended among all required days of the program minus holidays, calculated over the full duration of each participant’s enrollment in the program.
iii. Qualitative research: We conducted focus group discussions (FGDs) and in-depth interviews (IDIs) to explore participants lived experiences. These qualitative methods aimed to identify motivations for participation in the PRP, perceived benefits, and barriers to sustained engagement, with particular attention to gender-related factors influencing participation and psychosocial outcomes. Four FGDs (n = 28; males = 14, females = 14) were conducted across four participating childcare institutions. Each discussion group consisted of 6-10 participants of the same gender and age range (8-14 years or 15-19 years). FGDs were co-facilitated by gender-concordant youth investigators trained in focus group moderation, who ensured cultural and linguistic appropriateness. Simultaneously, we conducted four IDIs (n = 4; males = 2, females = 2) with current program implementers (Captains) who were also former *Positive Running* participants to further contextualize and deepen understanding of participant experiences. Sessions were conducted in English, Tamil, and Kannada, and lasted approximately 30-60 minutes and were conducted in a private setting to ensure privacy and confidentiality. With participants’ consent, discussions were audio-recorded, translated into English where necessary, transcribed, and de-identified prior to analysis.

### Ethics Statement

Ethical approval was obtained from the Institutional Review Boards of the Y.R. Gaitonde Centre for AIDS Research and Education (YRGCARE), Chennai, India, and the Johns Hopkins Bloomberg School of Public Health, Baltimore, USA. Informed consent, assent, and parental permission forms were available in both English and Kannada. Trained youth investigators obtained adult consent, youth assent, and parental/guardian permission, providing explanations in the participant’s preferred language to ensure comprehension and voluntary participation.

## Data Analysis

### Quantitative analysis

We analyzed sex-disaggregated sociodemographic characteristics (gender, age, viral suppression), intervention adherence, and prevalence of depression and generalized anxiety disorder participants (Table 1). Descriptive statistics were computed to summarize participant characteristics and key study variables. Using R (version 4.3.2), with statistical significance set at p <0.05 we analyzed data using the Fischer exact test to assess relationships between gender and key outcomes, including depression, generalized anxiety disorder, viral load suppression, and intervention adherence.^25^ Logistic regression models were used to estimate the unadjusted and age and sex-adjusted prevalence odds ratios of psychosocial health status in relation to program adherence; analyses were further stratified by sex to assess potential effect modification.

**Table 1.**
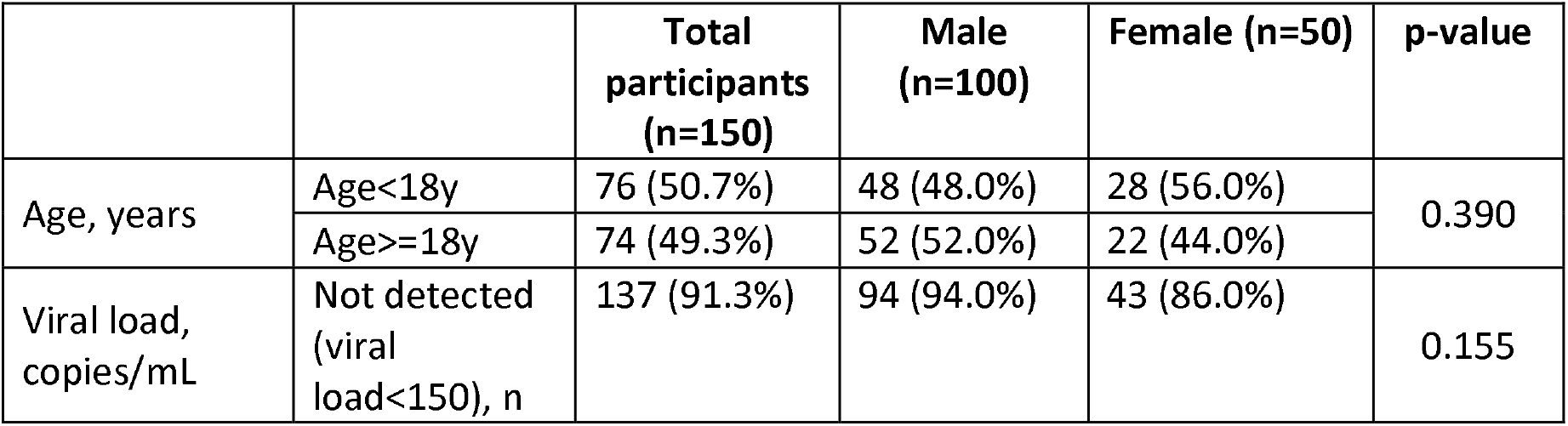
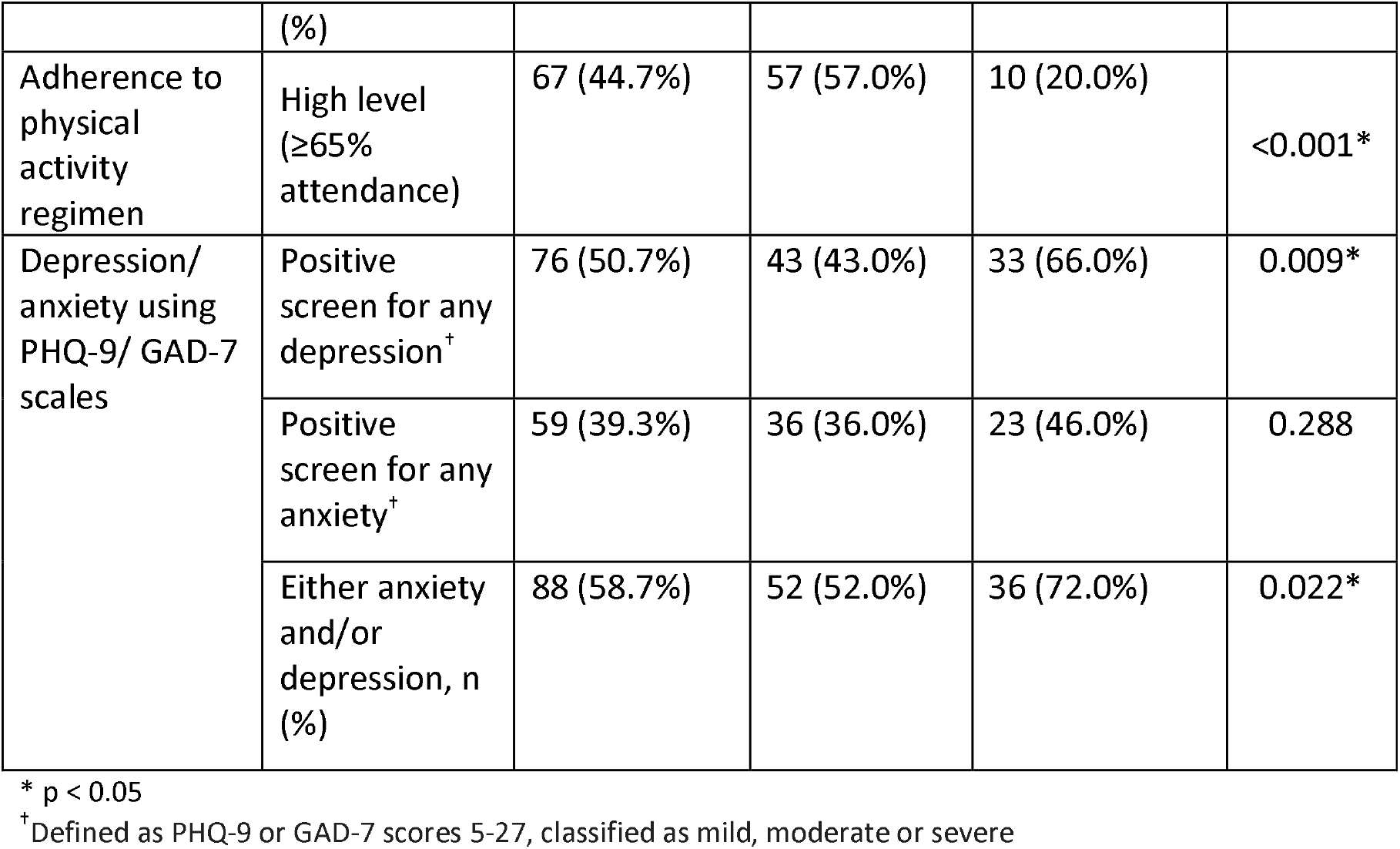
Sex-disaggregated sociodemographic characteristics (gender, age, viral suppression), intervention adherence, prevalence of depression and generalized anxiety disorder (GAD) among participants.

### Qualitative analysis

We analyzed data using grounded theory methodology and did thematic analysis guided by Braun and Clarke’s six-phase framework.^26^ Independent coders used iterative, constant-comparative coding to identify patterns and develop emerging themes grounded in participant responses. A vertical line-by-line review of the transcripts informed the development of an initial open-coding codebook, which was then consolidated and refined into an axial codebook. Both researchers independently cross-checked the application of axial codes, and intercoder reliability was established through discussion and consensus to ensure consistency in code interpretation and application.

### Theoretical frameworks

Through iterative analysis and reflexive dialogue, researchers identified emergent themes of gender-related barriers and facilitators that aligned with Intersectionality Theory and Self Determination Theory.^27,28^ The former contextualized how female participants’ overlapping marginalized identities including gender and HIV status compounded stigma and created multilayered barriers to intervention adherence.^27^ The Self Determination Theory that posits that health behavior motivation depends on fulfillment of three psychological needs: autonomy (perceived control over decisions), competence (self-efficacy in achieving outcomes), and relatedness (social connection and belonging), provided a framework for understanding mechanisms by which the intervention facilitated barrier mitigation.^28^ These two complementary frameworks were applied deductively to interpret participant narratives and ground emergent findings in established theory.

## RESULTS

### Survey results

Of the 172 participants who completed 12 months of the *Positive Running* intervention, 155 were children with HIV status confirmed prior to 10 years of age, and who were included in this analysis. Among these, 150 provided consent to participate in the survey. Reasons for non-consent included participant transfer (n=2), program discontinuation (n=2), and illness at the time of assessment (n=1). Median age of survey participants was 17 years [Interquartile range (IQR) = 15-19 years] with males constituting 66.7% of participants and females constituting 33.3%. Viral suppression (VL <150 copies/ml) was achieved by 91.3% of participants. Mean intervention adherence was 64.3% (females: 62%; males: 66%, SD = 10.69%), and was significantly lower among females than males, with only 20% of females achieving ‘high’ attendance (≥65%) compared to 57% of males (p < 0.001). (Table 1).

Over half of participants (58.7%) screened positive for at least one common mental health disorder (depression and/or anxiety); 50.7% screened positive for depressive symptoms on the PHQ-9, and 39.3% screened positive for anxiety on the GAD-7 scale (defined as mild and above category based on standard scoring procedures for PHQ-9 and GAD-7). Sex-disaggregated estimates revealed a significantly higher prevalence of depression and at least one common mental disorders among female participants compared with males. Specifically, 66% of females versus 43% of males met criteria for depression (p = 0.009), and 52% of females versus 72% of males met criteria for either/both depression and anxiety (p = 0.022). (Table 1).

Higher adherence to the intervention is associated with a lower likelihood of a common mental health disorder (unadjusted OR 0.44 (95% CI 0.23-0.85). When age-adjusted and stratified by gender, we found males with higher intervention adherence to have a lower likelihood of a common mental health disorder (aOR: 0.33, 95% CI: 0.14-0.75), although no association was found among females. (Table 2).

**Table 2.**
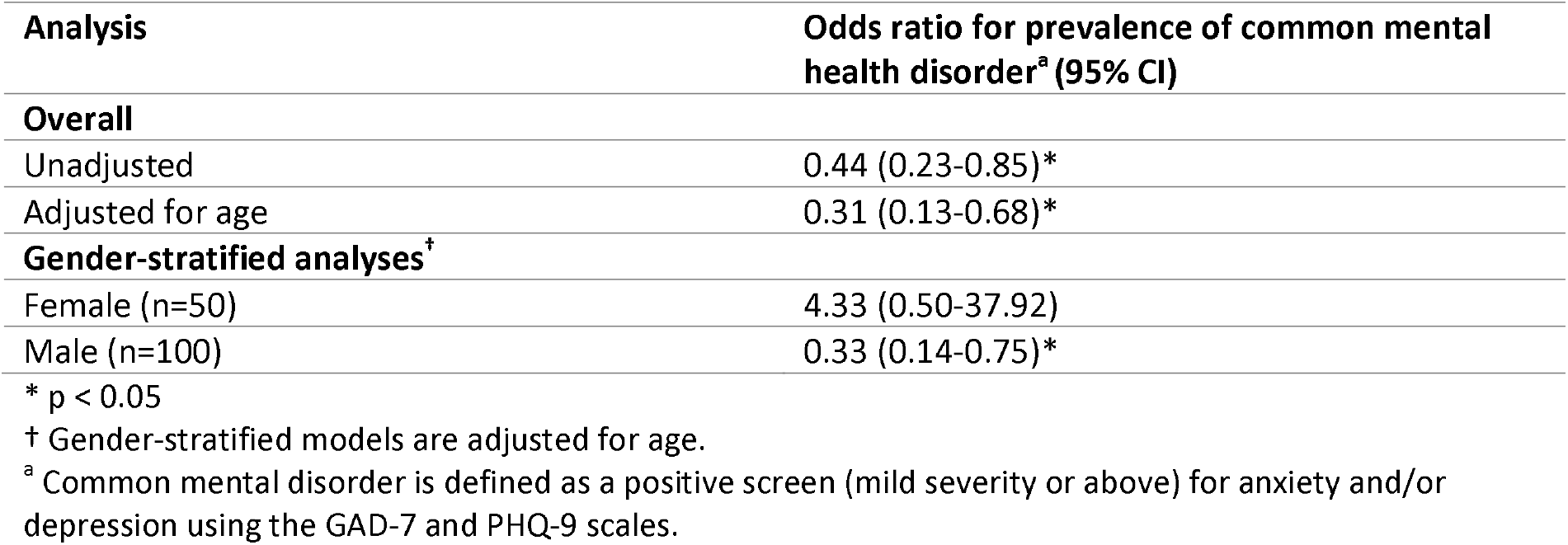
Association between intervention adherence and common mental disorder (reference category: low intervention adherence (≤65% attendance)

Compared with participants with low intervention adherence (≤65% attendance), those with higher adherence had significantly lower odds of screening positive for a common mental health disorder. In unadjusted analyses, higher adherence was associated with a 56% reduction in the odds of common mental disorder (OR = 0.44, 95% CI: 0.23-0.85). This association remained statistically significant and strengthened after adjustment for age (adjusted OR = 0.33, 95% CI: 0.15-0.71).

Gender was examined as an effect modifier; therefore gender-stratified estimates adjusted for age are presented. Higher adherence was associated with significantly lower odds of common mental disorder among males (OR = 0.33, 95% CI: 0.14-0.75), but was not statistically significant among females (OR = 4.33, 95% CI: 0.50-37.92), likely reflecting limited sample size and sparse outcome events among females. These findings suggest that the overall protective association between program adherence and mental health outcomes is driven primarily by male participants.

### Qualitative results

Thematic analyses from FGDs and IDIs explored the gendered, cultural, and structural factors shaping participants’ engagement in exercise. The first section explores barriers to female participation, revealing how intersecting vulnerabilities constrain their ability to engage in physical activity both within and outside institutional settings (Table 3). The second section examines facilitators of participation through the lens of self-determination, identifying how the *Positive Running* intervention fosters autonomy, competence, and relatedness among participants (Table 4).

**Table 3.**
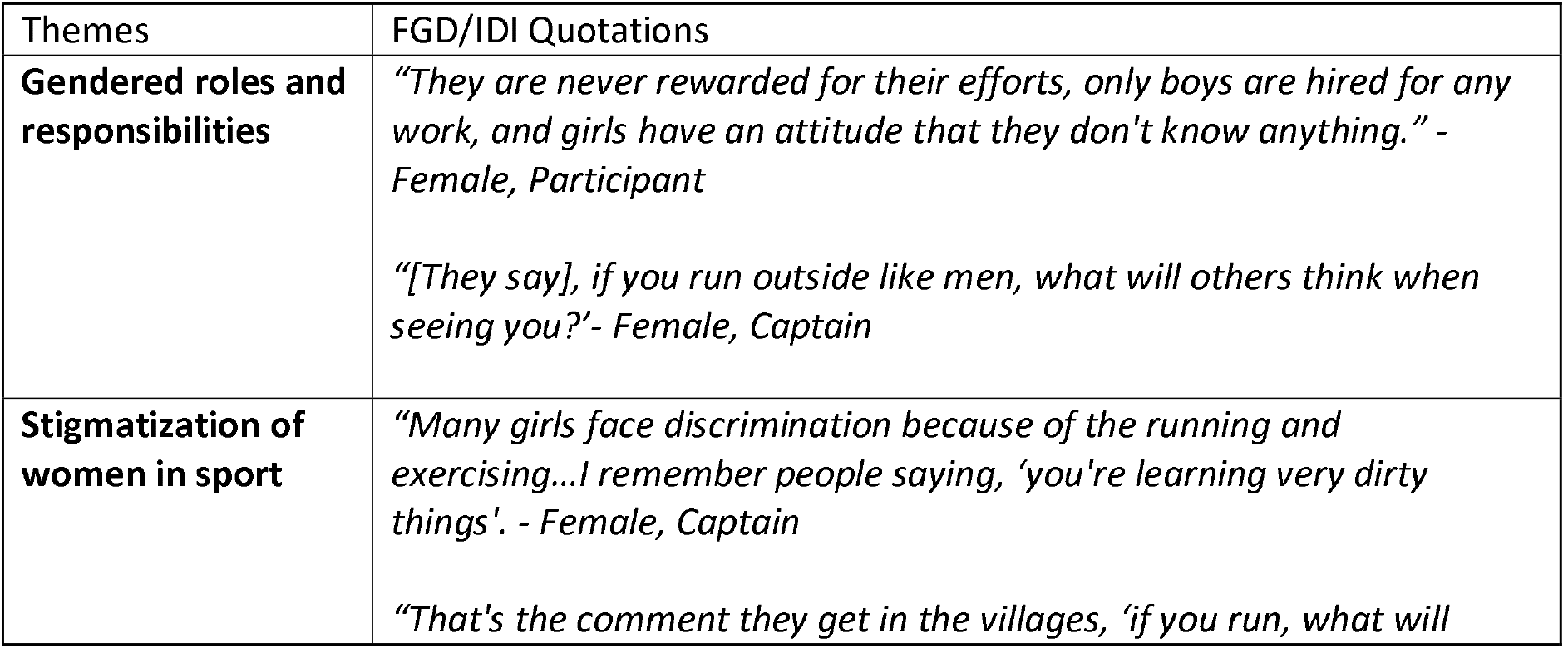
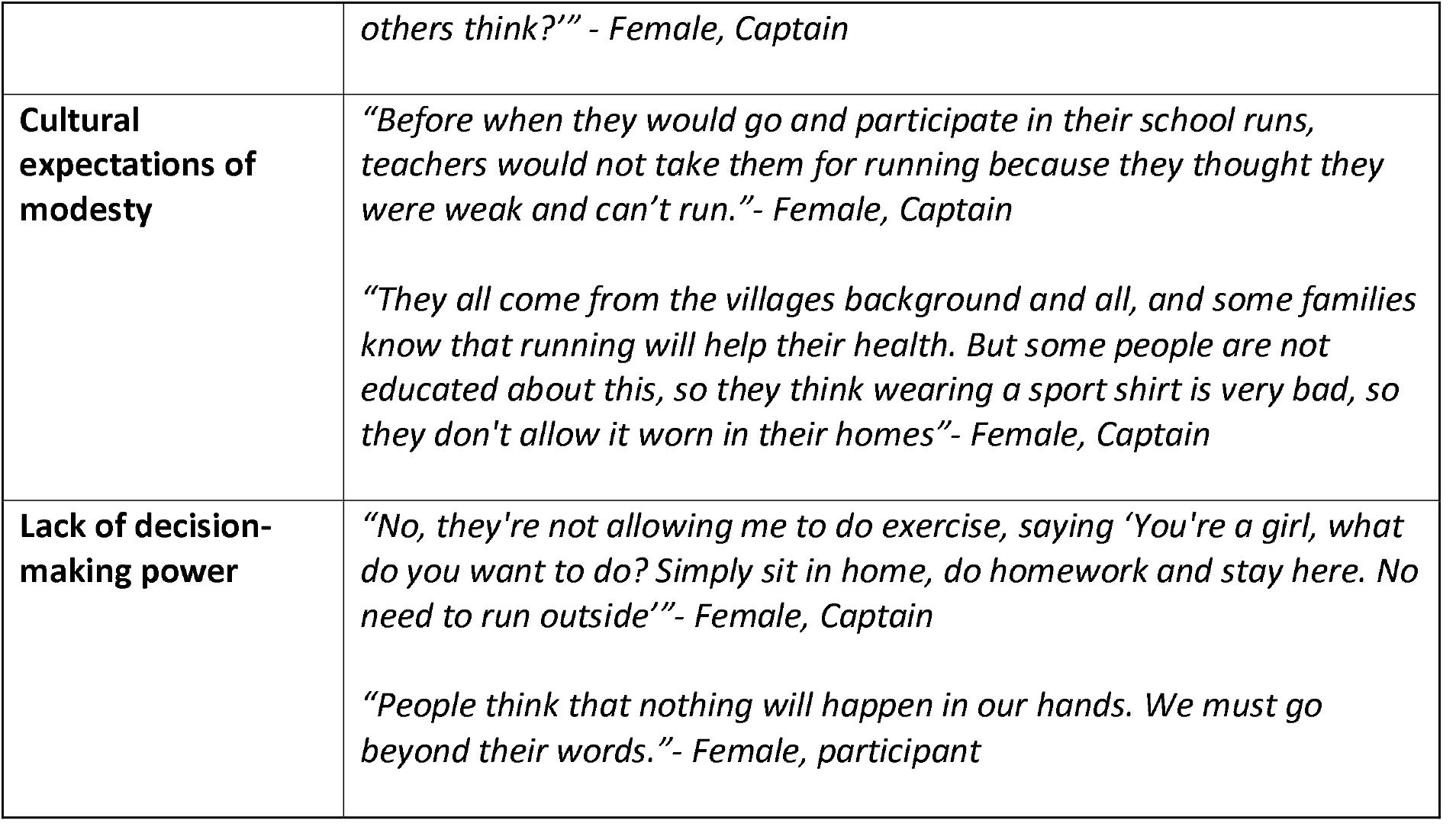
Themes of gender-related barriers contextualized using intersectional theory and participant quotations from focus group discussions and in-depth interviews.

**Table 4.** Themes of motivation contextualized using Self Determination Theory and participant key words and quotations from focus group discussions and in-depth interviews.

| Theme | Subthemes | Key Words | Illustrative Quote |
| --- | --- | --- | --- |
| **Autonomy** | Self-managed health<br>Leading and learning sessions during camps<br>Gender norms challenged with action | *“fit”*<br>*“strong”*<br>*“control”*<br>*“responsibility”*<br>*“equal”* | *“Many younger peers were so weak and having so much of cough and all. So, after they started running and exercising, it has reduced. So many have shared with me, ‘Before regularly I was getting cough and all, but after joining the run and doing running exercise my cough has reduced. I'm*<br>*becoming strong. I feel this strength.' They want to continue to be healthy, so they run."*<br>*- Female, Captain* |
| **Competency** | • Health knowledge<br>• Confidence built through distance milestones<br>• Recognition and achievement | *"improvement"*<br>*"achievement"*<br>*"capable"*<br>*"goals"*<br>*"better"* | *"When they started, they were not doing much. Most of the peers were telling me that it's difficult to run saying, 'I cannot run'. Then, we slowly pushed them to do 2 kilometers, then 5 kilometers. Then they say, 'we feel that we can do more than before, like we can do better, more than other children... because some children don't get opportunity to do, but we get to."*<br>*- Female, Captain* |
| **Relatedness** | • Peer bonding through team runs<br>• Collective motivation through Captains' mentorship<br>• Increased sense of belonging fostered through mentoring and being mentored | *"share"*<br>*"together"*<br>*"team"*<br>*"inspire"*<br>*"encourage"* | *"Like if I run alone, I don't feel any motivation because I'm all by myself. But when we come together for the camps, we get many friends. We have lots of fun. We all will jump in and we will have lots of fun. So that's the key motivation I think."*<br>*- Male, Captain* |

#### I. Barriers to girls’ participation in physical activity

Qualitative findings revealed insights on how intersecting challenges consisting of gender roles and responsibilities, stigmatization of women in sport, cultural expectations of modesty, and lack of decision-making power, limited female participation (Table 3).

##### Gendered roles and responsibilities

Female participants shared that when they attempted to exercise either within or outside of the institution, they often encountered negative responses from both family and community members.

*“So the parents feels that our girls have to be a certain way only and they should not do boys’ work. Girls can only do girl work and homework, so they should not do running and other things. They have to be inside the home and work.” - Male, Captain*

Such responses reflected prevailing gender norms that view physical activity and outdoor exercise as inappropriate for girls, who are expected instead to devote their time to traditional household responsibilities, or “girl’s work”. These external perceptions that exercise is a “man’s activity” reinforce gender norms that discourage female participation in physical activity.

##### Stigmatization of women in sport

Participant responses revealed a common theme of gendered stigma surrounding women engaging in sports/physical activity. This injunctive norm often took an active form in discrimination by neighbors and community members outside of the institution, viewing women exercising as “dirty” resulting in decreased participation due to fear of social judgement and ostracization. This stigmatization served as a significant demotivating factor for female participants.

*“That’s the one thing; they talk bad about the girls. Because of that, they stop running or they don’t have any interest or confidence to do it” - Female, Captain*

##### Cultural expectations of modesty/femininity

A barrier to continued participation at the community level centered on commonly held cultural beliefs that women should strive to attain modesty and femininity in the way they present themselves in public. For female participants, sportswear that is ideal for running such as a shirt and pants were often met with criticism from community members and disallowed by family members upon exiting the house to run/exercise. One participant shared that running in their village felt restrictive, driven by fears of community scrutiny and negative judgments about their upbringing.

*“We feel that we have done something wrong. No one is interested in doing it again if they say no. What will [they] talk again, what will [they] say?”- Female, participant*

##### Lack of decision-making power

Due to their intersectional identity as both women and young adults living with HIV, participants often expressed a lack of decision-making power in terms of being able to exercise outside of the institution. In a patriarchal society where males are the head of the household, (the father is the head before marriage and the husband assumes the role post-marriage) the decision to leave the household is often placed solely on the male figurehead. Therefore, despite personal motivation, girls are often unable to independently decide to continue exercising due to social and familial constraints.

*“We see in many villages there are many families they don’t send their girls anywhere outside, especially to work or study in different places apart from their village. They won’t allow girls, or they won’t encourage them to run.”- Female, Captain*

#### II. Facilitators of girls’ participation in physical activity

The qualitative findings illuminate how gender-related components of autonomy (control over health and wellness, independence), competency (exercise/nutrition knowledge, self-efficacy/empowerment), and relatedness (peer support networks, challenging gendered norms) from the Self-Determination Theory has motivated participants to continue sustained physical activity, as well as enhanced motivation in other key life areas including self-confidence and educational attainment. (Table 4).

#### Autonomy

Participants expressed how sustained physical activity and educational sessions on health and hygiene practices has allowed them to gain greater control over several dimensions of their overall wellness. Youth were able to exercise choice, develop self-regulation, and engage in health-promoting behaviors driven by intrinsic motivation rather than external influence. Decreased instances of illness increased emotional control, and experiences of joy/happiness related to participation were common testimonies.

*“Last year there was no exercise, so I fell sick two or three times, this time I did not fall sick even once because of running and exercise*.*” -* Male, participant

Additionally, female Captains shared experiences of increased independence including ability to travel further distances, financial self-sufficiency and greater resilience because of participation.

*“I feel more confident in me because I have learned so many things. Now it may be travelling, it may be spending money. It may be training others, talking to others, like giving them suggestions. It has improved many things in my life, and I can also take up many responsibilities. It has mainly increased my confidence and especially increased my confidence in myself. Now I can trust myself so much*.*”* - Female, Captain

Moreover, when it came to challenging gender norms within their own communities, female participants felt more empowered to take independent action against restrictive expectations.

*“Let them think what they want, we have to take care of our own health matters*.*” -* Female, participant

#### Competency

The theme of competence captures participants’ growing confidence in their abilities, skills, and self-efficacy as they engaged in structured physical activity and health-promoting behaviors. As a result of the intervention’s nutritional and general health/hygiene trainings, participants expressed greater knowledge and an ability to support themselves and other peers when experiencing difficulties related to these topics.

*“We know that when we are running and exercising, we need to have good nutritious food daily. Like if we are getting groggy, to eat more egg or any fruits and vegetables. So, we have to have our daily nutritious food on time and mainly take your tablets on time*.*”-* Female, Captain

Furthermore, setting goals such as completing selected regional runs and marathons increased self-efficacy (the belief in oneself to overcome challenges and achieve goals). These mastery experiences as well as reported positive emotional changes resulting from sustained participation (increased focus, confidence) had an empowering effect in other life areas for participants including increased engagement in school and setting career goals.

*“There are any running competitions in our school, if we run well here and practice well, we can go there and run well. If we run elsewhere, our school will get a good name and our teacher will also be happy*.*”* - Female, participant

#### Relatedness

The theme of relatedness captures participants’ sense of belonging and mutual support, highlighting how shared experiences and peer relationships engendered social growth. Participants described how programmatic activities including running camps that are part of the intervention created an enabling environment for participants to create sustainable friendships.

*“Running with new people and participating and talking with new people, it’s very nice. We can share how we are feeling while running. What are our difficulties. We can share all those things. All are challenges about running while we are running and all. So, it will be good to share each other’s opinion. So, we can get to know each other better and we can keep in contact with them*.*” -* Female, participant

Additionally, presence of Captains during institutional visits provided participants an opportunity to share their concerns through peer-counseling and provided motivation for participants to continue adherence when experiencing gender-related barriers.

*“They see me, and they’ll also start doing and they’ll be also saying, ‘I want to be like you, I want to do this exercise like you’ and they say, ‘How are you doing these many things?’. So I say that I learned from the others and I’m teaching for them. Then I say, ‘you have to learn from me, and you have to teach for others’”. -* Male, Captain

*“Courage comes when [Captain] is with us”* - Female, participant

The *Positive Running* intervention’s approach in bringing together female and male participants to achieve similar fitness goals produced a change in the perception of male participants regarding female abilities, an important step towards gender equality. Male participants recognized and accepted female participation and expressed support for their female peers engaging alongside them in sports and games.

*“Nowadays boys and girls are at different institutions. So, when they come back together for a camp, they can see each other’s capabilities. Like sometimes boys may be thinking that, ‘Yeah, we are good at something’ and they may not know that there’s a girl better than them at it. So, when they come together, they can understand other’s abilities so that they can be inspired and they can change their mindset that girls can do it too*.*”*-Female, Captain

*“I want them [girls] to run forward like us*.*” -* Male, participant

Male participants described how seeing girls excelling in sports challenged preconceived, socially constructed differences in abilities across genders (i.e. girls can’t run as far or as fast, boys lead physical activities while girls only socialize). Female participants expressed how visible female role models (female Captains) encouraged their belief in pursuing and achieving the same goals as their male counterparts, fostering an equitable environment of inclusion and encouragement.

*“The program is showing even the girls can run 5K and 10K. It is an example for the girls that you can also play any games, and you are equal to the boys*.*”-* Male, Captain

## DISCUSSION

This mixed-methods evaluation demonstrates that the *Positive Running* intervention’s gendered impact on health and psychosocial factors among adolescents and young adults living with perinatally acquired HIV. The integration of structured exercise, mentorship, and peer leadership suggested beneficial changes in self-determination particularly among female participants who face intersecting barriers of stigma, gender norms, and limited agency.

### Health and psychosocial gains

Structured physical activity participation among people living with HIV has been shown to enhance well-being through the interaction of physiological, psychological, and social mechanisms.^29,30^ In our study, despite moderate intervention adherence (64%), participation was consistent enough to generate meaningful psychosocial benefits. The biochemical basis for these effects is well established; exercise regulates neuroendocrine pathways such as cortisol and stimulates the release of endorphins, serotonin, and brain-derived neurotrophic factor, which are neurochemicals linked to improved mood, cognitive functioning, and stress resilience.^31,32^ Similarly, psychological benefits emerge through the cultivation of mastery, goal attainment, and self-efficacy, which reinforce autonomy and competence, core components of Self-Determination Theory.^28,29^ These benefits have been predominantly documented in adults living with HIV; notably, qualitative narratives from our study revealed comparable self-reported gains among youth with HIV, including reductions in fatigue, greater emotional control, and heightened self-esteem. These findings highlight the potential of low-cost, community-based physical activity programs to complement biomedical HIV care.^33–35^

### Peer-led model and the pathway to empowerment

According to WHO estimates, steep increases in insufficient physical activity between 2000 and 2022 were observed in the high-income Asia Pacific and South Asia regions, with trends projected to continue rising through 2030.^36^ The *Positive Running* model’s distinctive peer-led structure emerged as a crucial enabler of sustained engagement and empowerment. Captains, who are trained peers with lived experience in the intervention, served as relatable mentors who modeled self-efficacy and normalized physical activity. Through shared experiences, participants reported increased autonomy (“*I can trust myself so much*”), competency (“*we know how to eat and exercise properly*”), and relatedness (“*when we come together, we get motivation*”), illustrating how the intervention strengthens the three pillars of self-determination. This alignment suggests that peer-led, participatory interventions may activate intrinsic motivation, leading to durable behavior change and psychosocial growth. While these mechanisms promote immediate improvements in emotional regulation and well-being, they also underpin sustained adherence to care and long-term psychosocial resilience,^37^ reinforcing the value of peer-led model of physical activity and community-building as a critical active ingredient in a mental health intervention.

### Gender-transformative approach

In India, deep rooted expectations around modesty, safety, and domestic responsibilities restrict women’s mobility and ability to participate in physical activity.^38,39^ This is embedded in the perception of physical activity as a symbol of undesirable modernity, a rejection of traditional values that can invite social disapproval and harassment, to which young women are particularly vulnerable.^38,39^ Even when women succeed in navigating these sociocultural barriers, they continue to face gender bias in the allocation of funding, sports infrastructure, and training opportunities, which disproportionately favor men.^38^

These structural inequities are reflected in gendered differences in mental health outcomes, particularly the threefold higher odds of depression and/or anxiety among female participants. ^40^ Prior research across diverse cultural contexts, reinforced by findings from this study’s focus group discussions, reveals that women’s participation in sport is constrained across four interconnected domains: physiological, psychological, sociocultural, and environmental.^40,41^ In the Indian context, female participants’ accounts of community disapproval, poor autonomy, and cultural norms surrounding femininity were more prominent, revealing that even within institutional settings, internalized gender expectations limit engagement. These findings align with and extend existing literature on gender barriers to physical activity by demonstrating how structural and cultural factors interact to constrain girls’ opportunities.^38,39,42,43^ For example, prior studies describe how adolescent girls’ physical mobility is curtailed by social surveillance and moral policing, both of which intersect with the stigma surrounding HIV.^18,44^ The identification of lack of decision-making power as a barrier (Table 3) is particularly noteworthy, as it highlights how adolescent girls often lack agency in determining their own physical activity participation. This power asymmetry operates at multiple levels, within families, educational institutions, and broader community structures.

The *Positive Running* intervention appears to mitigate several gender-based barriers to physical activity by addressing constraints across physiological, psychological, sociocultural, and environmental domains. By offering structured and progressive exercise sessions offered under supervision, the intervention reduces fears related to injury and fatigue, thereby increasing participants’ confidence in their physical capacity. The peer-led and non-competitive nature of the program helps alleviate psychological barriers by creating an affirmative environment that minimizes judgment on wearing athletic clothing and frames physical activity as collective participation. The *Positive Running* intervention attempts to provide a socially sanctioned and gender-affirming space for exercise, helping legitimize girls’ participation within communities where female mobility and sport engagement are often constrained by norms of modesty and HIV-related stigma.

The differential association between intervention adherence and mental health outcomes for males and females merits further discussion. Our finding that male participants showed a stronger association between intervention adherence and improved mental health status compared to female participants may reflect gendered and intersectional pathways shaping psychological distress among adolescents born with HIV. Adolescent girls and young women with HIV often face more profound social and structural stressors, including stigma, poor autonomy, caregiving burdens, and exposure to violence, that may attenuate the mental health benefits of adherence-focused interventions, such that consistent participation alone is insufficient to reduce symptoms of anxiety or depression. From a self-determination theory perspective, adherence may also represent qualitatively different motivational constructs across males and females. Among male youth, higher adherence may reflect greater autonomy and perceived competence, thereby translating participation into measurable mental health benefit. In contrast, adherence among female youth may be less self-directed, and more strongly shaped by external regulation or obligation, particularly in contexts where mobility, participation, and health-related decision-making are constrained. In such cases, adherence may not satisfy autonomy needs and may therefore confer limited mental health protection despite high attendance. These findings underscore the importance of gender-responsive intervention design for adolescents born with HIV, in which strategies to promote participation are complemented by components that explicitly address structural stressors and support autonomous engagement for females. The *Positive Running* intervention may benefit from a deeper integration of psychosocial support, challenging of social norms, and agency-enhancing elements in order to ensure that adherence translates into mental health benefit across genders.

### Theory informed explanatory model for the Positive Running intervention

Figure 1 illustrates how *Positive Running* addresses the intersecting vulnerabilities of adolescent girls living with HIV through a framework informed by Intersectionality Theory and grounded in Self Determination Theory.^27,28^ Although not explicitly explored in this study, the findings underscore the importance of recognizing the multiple, overlapping identities of participants, including being of young age, female, living with HIV, and often socioeconomically disadvantaged or orphaned, which together heighten vulnerability and compound risks of poor health outcomes. These intersecting factors can intensify barriers to participation, underscoring how gendered and structural inequities shape health experiences. As shown in Figure 1, program components promoting autonomy, competence, and relatedness, through self-led fitness activities, skill-building exercises, and peer mentorship, enabled participants to build confidence, self-management, and intrinsic motivation. Collectively, these elements helped reframe traditional gender expectations, strengthen self-efficacy, and promote more equitable engagement for female participants.

**Figure 1.**
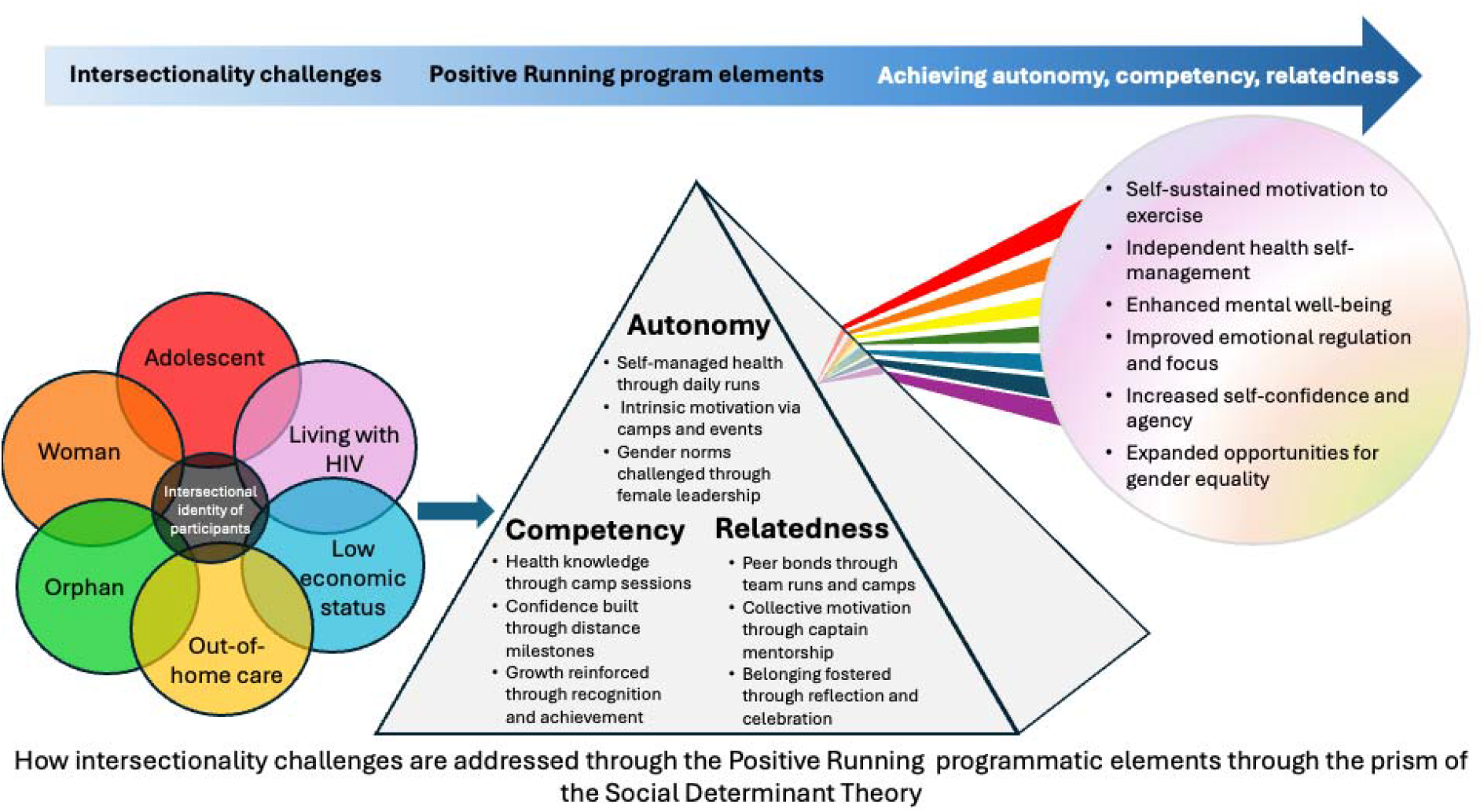
Intersectional Theory and Self Determination Theory applied to female participant experiences

Findings from *Positive Running* reinforce the need for adolescent-centered HIV programs that integrate physical activity, mental health, and gender equity into a cohesive framework. While biomedical adherence and viral suppression remain critical, interventions that nurture autonomy, knowledge-building, and peer connectedness may enhance long-term resilience and retention in care. The peer-led approach is potentially both scalable and sustainable, requiring minimal resources while promoting local ownership and continuity. Scaling this model across community and institutional settings could strengthen India’s adolescent HIV response, particularly for girls who remain underserved within conventional health systems.

### Strengths and limitations

This study’s mixed-methods design represents a key strength, enabling triangulation of quantitative mental health and participation data with rich qualitative insights about barriers and facilitators. The sex-disaggregated analysis provides important nuance often missing from adolescent physical activity and HIV research. The use of a community-based participatory approach, centering the study implementation with investigators with lived experience, ensured that the research process remained participatory and sensitive to the lived experiences of youth with HIV.

Several limitations warrant consideration. The mental health screening tools (PHQ-9, GAD-7), while validated, represent screening rather than diagnostic assessments. The cross-sectional nature of the mental health data precludes causal inference about relationships between physical activity participation and mental health outcomes. The qualitative sample, while providing rich insights, did not capture mental health outcomes. Additionally, pre-existing mental health comorbidities may have influenced patterns of physical activity adherence whereby youth with less mental health distress were better able to engage with the intervention. Data were primarily self-reported, which may introduce social desirability or recall bias. The study’s single geographic focus and limited sample size restrict generalizability of the findings to other settings, though the identified barriers are consistent with findings from diverse cultural contexts.^42^

## CONCLUSION

The *Positive Running* intervention demonstrates that community-based, peer-led physical activity is not only feasible but has the potential to be transformative for youth living with chronic health conditions. Findings underscore the importance of sustained investment in, gender-responsive, community-embedded physical activity interventions in South Asian contexts where females face intersecting structural, social, and health-related vulnerabilities. Future research should prioritize rigorous evaluation of the long-term effects of such interventions on physical, mental, and psychosocial health outcomes, as well as their cost-effectiveness and scalability. Given its adaptable design and emphasis on peer leadership, autonomy, and empowerment, the *Positive Running* model offers a promising framework for extension to other adolescent populations with chronic conditions. Integrating physical activity-based, community-led approaches into broader adolescent health strategies may represent a scalable pathway to promote resilience, agency, and well-being across diverse settings.

## Data Availability

All data produced in the present study are available upon reasonable request to the authors

